# Improving irregular temporal modeling by integrating synthetic data to the electronic medical record using conditional GANs: a case study of fluid overload prediction in the intensive care unit

**DOI:** 10.1101/2023.06.20.23291680

**Authors:** Alireza Rafiei, Milad Ghiasi Rad, Andrea Sikora, Rishikesan Kamaleswaran

## Abstract

**Objective:** The challenge of irregular temporal data, which is particularly prominent for medication use in the critically ill, limits the performance of predictive models. The purpose of this evaluation was to pilot test integrating synthetic data within an existing dataset of complex medication data to improve machine learning model prediction of fluid overload.

**Materials and Methods:** This retrospective cohort study evaluated patients admitted to an ICU *≥*72 hours. Four machine learning algorithms to predict fluid overload after 48-72 hours of ICU admission were developed using the original dataset. Then, two distinct synthetic data generation methodologies (synthetic minority over-sampling technique (SMOTE) and conditional tabular generative adversarial network (CT-GAN)) were used to create synthetic data. Finally, a stacking ensemble technique designed to train a meta-learner was established. Models underwent training in three scenarios of varying qualities and quantities of datasets.

**Results:** Training machine learning algorithms on the combined synthetic and original dataset overall increased the performance of the predictive models compared to training on the original dataset. The highest performing model was the metamodel trained on the combined dataset with 0.83 AUROC while it managed to significantly enhance the sensitivity across different training scenarios.

**Discussion:** The integration of synthetically generated data is the first time such methods have been applied to ICU medication data and offers a promising solution to enhance the performance of machine learning models for fluid overload, which may be translated to other ICU outcomes. A meta-learner was able to make a trade-off between different performance metrics and improve the ability to identify the minority class.

## Introduction

Medication regimens in the intensive care unit (ICU) are notoriously complex, and meaningful analysis poses a unique challenge to clinicians at the bedside and Big Data scientists alike [1-5]. A unique element of medication data includes the degree of granularity necessary for appropriate interpretation (e.g., drug name, dose, frequency, formulation, etc.) [6]. A result of this complexity is that limited attempts have been made to incorporate the entire medication regimen into prediction modeling despite medication therapy’s essential role in patient outcomes [7].

Some initial studies have shown promise, including improved mortality prediction with the incorporation of medication data and machine learning [1] as well as the discovery of novel pharmacophenotypes associated with patient outcomes, particularly with the use of a novel common data model to make medication data more machine-readable [6 8]. However, the efficacy of these advanced technologies is contingent upon the availability and quality of data for model training [9]. In many cases, progress is hindered by datasets that are insufficient, imbalanced, and biased [10 11]. Moreover, the sensitive nature of health-related information coupled with strict regulatory and security requirements complicates the acquisition and usage of healthcare data for machine learning model development [12]. Given the complexity of medication data, alternative strategies to enrich datasets are paramount to fully leverage artificial intelligence in this domain.

Synthetic data generation has emerged as a potential solution to bypass constraints associated with real-world healthcare data [13-15] but to date, the methodology has not been applied to the ICU medication domain. New data are generated by replicating real-world data’s essential characteristics while also adding variation, which can facilitate the development of robust and reliable machine learning models [16 17]. The adoption of synthetic data in the healthcare domain may alleviate limitations associated with protected health information and can help address data quality, completeness, and representation issues by generating balanced and comprehensive datasets that reduce biases and improve the generalizability of machine learning models [18 19]. However, synthetic data generation methods for critically ill patients and more specifically for evaluation of medication regimens of intensive care unit (ICU) patients are still in the nascent stages of development.

Given the complexity of ICU patient management, a specific use case for predicting fluid overload was developed for initial synthetic data generation. Fluid overload is a common though often unintentional consequence of caring for critically ill adults in the initial phase of their ICU stay. Despite the clinical need for volume resuscitation or intravenous (IV) medications that can lead to fluid overload, it is associated with increased rates of ICU complications, including acute kidney injury, use of invasive positive pressure ventilation, and prolonged ICU stay [20-23]. As such, fluid management is a clinically complex challenge. The ability to predict patients at risk of fluid overload and its sequelae has the potential to improve the implementation of fluid stewardship initiatives. To date, studies focused on predicting fluid overload using artificial intelligence-based approaches are limited [24]. Sikora et al. [25] contrasted the efficacy of the logistic regression model against traditional machine learning methods for fluid overload prediction. Although machine learning models outperformed logistic regression, they struggled to accurately predict positive cases of fluid overload (potentially due to an imbalance of fluid overload events in the original dataset). Despite these limitations, this study was the first to take into account full medication regimen data in addition to patient specific data to enhance predictive abilities.

The present study sought to build on this concept of incorporating comprehensive medication data as well as other patient specific data to predict fluid overload within 48-72 hours post-ICU admission by applying synthetic data generation methods. The main contributions of this study include: (1) employment of two synthetic data generators (synthetic minority over-sampling technique (SMOTE) and conditional tabular generative adversarial network (CTGAN)) to augment the available training data by mimicking real-world data characteristics and oversampling positive fluid overload cases to address the limitations posed by the quantity and quality of the original dataset; (2) Developing a generalizable and interpretable meta-learner to address the limitations of other machine learning models when predicting positive fluid overload cases.

## Materials and methods

### Dataset

De-identified data from the University of North Carolina Health System electronic medical record (EMR) data (Epic Systems, Verona, WI) housed in the Carolina Data Warehouse (CDW) were extracted by a trained CDW data analyst. In this process, a random list of 1,000 patients was generated between October 2015 – October 2020. From here, patients on their index ICU admission that had fluid balance data for the first 72 hours available were included.

The protocol for this study was reviewed and approved by the Institutional Review Board (IRB) of record at the University of Georgia (approval number: (PROJECT00002652); approval date: October 2021). Due to the retrospective, observational design, waivers of informed consent and HIPAA authorization were granted. The procedures followed in the study were in accordance with the ethical standards of the IRB and the Helsinki Declaration of 1975 [26]. Study reporting adheres to the STrengthening and reporting of OBservational data in the Epidemiology statement [27].

This was a retrospective, observational study of adults admitted to the ICU assessing the primary outcome of fluid overload at 48-72 hours (i.e., day 3) following ICU admission. The definition of fluid overload was a positive fluid balance in milliliters (mL) greater than or equal to 10% of the patient’s admission body weight in kilograms (kg) [20 28]. Predictor variables included four categories: 1) ICU baseline: age *≥*65 years, sex, admission to a medical ICU, primary admission diagnosis category (including cardiac, chronic kidney disease, rhabdomyolysis, heart failure, cirrhosis, pulmonary arterial hypertension, hepatic, pulmonary, pancreatitis, sepsis, trauma, other), and select co-morbidities of chronic kidney disease or heart failure; 2) 24 hours after ICU admission: APACHE II and SOFA score, use of supportive care devices including renal replacement therapy and invasive mechanical ventilation, serum laboratory values including albumin <3 mg/dL, bicarbonate <22 mEq/L or >29 mEq/L, chloride ≥110 mEq/L, creatinine ≥1.5 mg/dL, lactate ≥2 mmol/L, potassium ≥5.5 mEq/L, sodium ≥148 mEq/L or <134 mEq/L, fluid balance, and presence of acute kidney injury (as defined by need for renal replacement therapy or serum creatinine greater than or equal two times baseline); 3) Medications at 24 hours: MRC-ICU, vasopressor use in the first 24 hours, use of continuous medication infusions, and the number of continuous medication infusions.

### Model development workflow

The study’s workflow is summarized in Figure 1, beginning with data collation and concluding with the final prediction. The collated ICU data initially went through a data processing stage. In this step, any variables associated with missing proportions exceeding 30% were omitted, except for the SOFA score. Afterward, the multiple imputation by chained equations (MICE) technique was used to handle complex missing data patterns and different variable types, while considering the uncertainty around imputed values [29]. Linear regression was used for the imputation of continuous variables, including SOFA, APACHE II, fluid balance (mL), and the amount of fluid overload. Logistic regression was adopted during the imputation process for binary variables while polytomous logistic regression was used for multi-level variables. Next, the processed data was fed into a generator to produce synthetic data. Simultaneously, the SMOTE technique was applied for oversampling the processed dataset. These synthetically generated datasets were combined with the original dataset and introduced to the discriminator. Of note, all of the synthetically generated data were used exclusively for training purposes. Ultimately, the data passed through a discriminator, which was a machine learning model to deliver the final prediction for fluid overload in ICU patients.

**Fig. 1.**
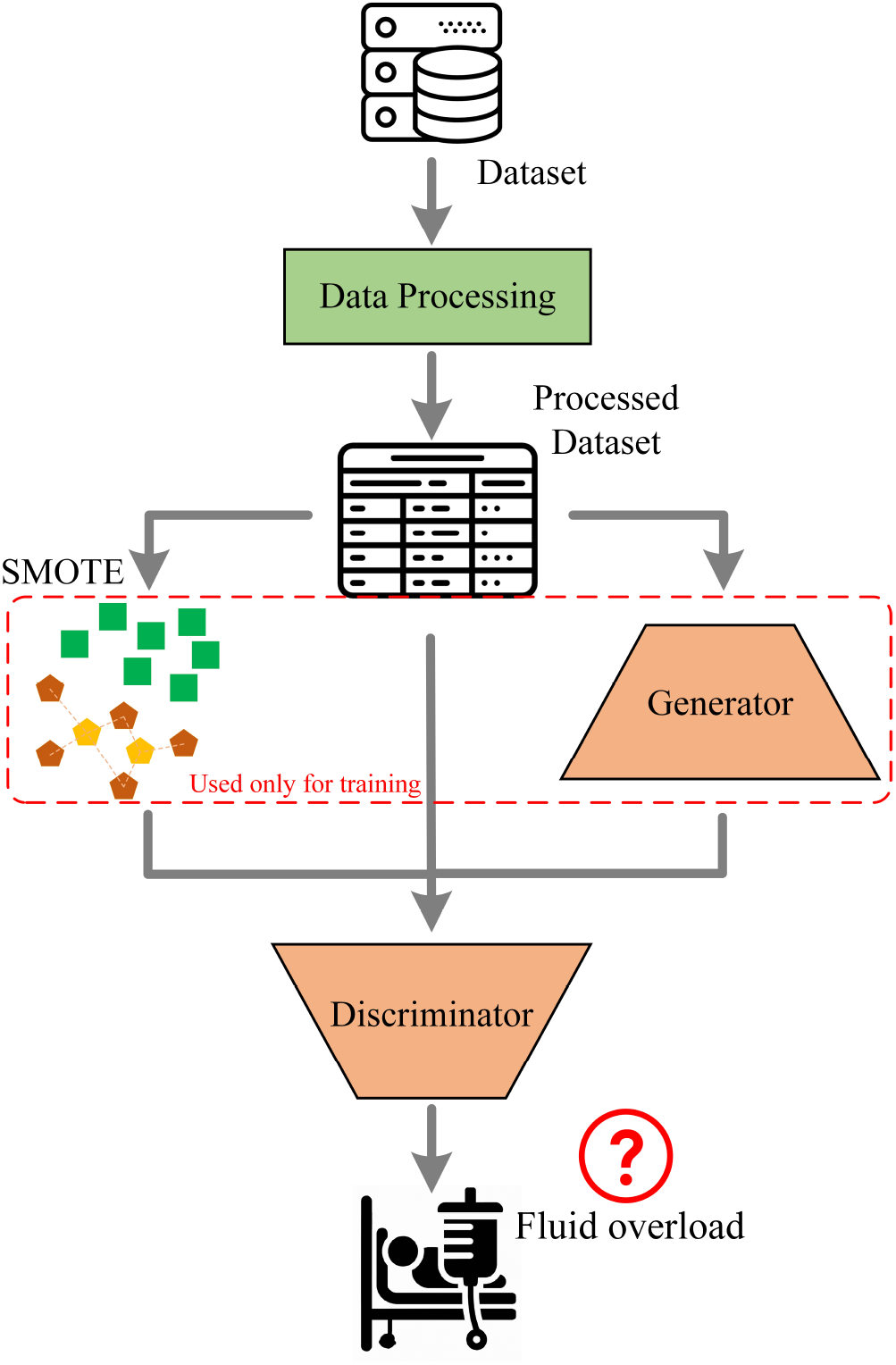
The workflow of predicting fluid overload.

### Synthetic data generation

The collected dataset was limited by an imbalance in the derived fluid overload labels with only 10% of the cases being labeled as positive for fluid overload. This skewness may create bias in the prospective models trained on the dataset predisposing them to focus primarily on the majority class while disregarding the minority class. This inherent bias potentially leads to the deceptive performance of the models: they might display high accuracy rates, but their ability to identify the positive cases would be considerably undermined. Given that the primary objective of the classifiers under development is to effectively identify patients with fluid overload, this limitation is notable.

In general, upper, lower, or hybrid sampling methods are the most common approaches to bring balance into the classification datasets. SMOTE is a widely employed technique for addressing the class imbalance in datasets and is particularly effective when the minority class instances are insufficient [30]. Instead of merely replicating the minority class samples to augment their representation, SMOTE generates synthetic instances based on the variable space similarities between existing minority samples. This method promotes diversity within the minority class helping to improve the model’s ability to generalize and thereby potentially enhancing its performance in predicting minority class instances. In this study, we assessed the potential influence of SMOTE-generated synthetic instances of positive fluid overload cases on the predictive performance of machine learning models. For this aim, synthetic instances were generated by selecting existing instances and creating new EMRs between these instances and their nearest neighbors. This process was continued until generating 777 fluid overload positive cases to balance the distribution of the labels within the dataset.

The second synthetic dataset was generated using a CTGAN model with the exact size of the original dataset but a more balanced distribution in terms of labels (44% positive cases). The purpose of this dataset was to provide more data in the training process to help machine learning algorithms decipher inherent patterns in the data and elevate the balance between the binary class of negative and positive fluid overload cases. CTGAN employs a conditional variant of the GAN model that allows for the generation of synthetic data with certain specified characteristics. This conditional aspect makes the model able to produce data samples that adhere to specific conditions. CTGAN can learn the complex and nonlinear interdependencies between different variables in the data, can be highly effective in non-Gaussian distributions, and can manage both categorical and continuous variables and discrete and continuous distributions. The rationale for CTGAN was to substantially enhance the volume and diversity of the training set by generating a balanced synthetic dataset with regard to label distribution. The CTGAN model was developed with previously proposed architecture [31]. This model created a dataset identical in terms of the number of features and instances as the original dataset yet maintained approximately the same distribution of positive and negative labels. This enhanced dataset was then incorporated with the original dataset for training the machine learning models. By enriching the training data, this approach ideally not only offers additional training samples for machine learning models but also increases the number of whole positive fluid overload cases to enhance the generalization robustness of the learning models.

### Machine learning algorithms

Four different machine learning approaches were employed to predict fluid overload at the first 72 hours of ICU admission: logistic regression (LR), support vector machine (SVM), XGBoost (XGB), and random forest (RF). Using the original dataset, these models did not demonstrate appropriate performance, particularly for detecting positive cases (sensitivity) [25]. Thus, in addition to the synthetic data generators, a meta-model was incorporated into the original study design to bolster the efficacy of the predictive fluid overload model. This approach led to the creation of a stacking ensemble model designed to train a meta-learner for the prediction of fluid overload. The fundamental methodology is rooted in the concept of stacked generalization, which leverages the combined predictive power of multiple models. It operates by generating individual predictions from an array of models. These predictions are then consolidated by a meta-model, yielding a holistic final prediction. The essence of this technique lies in the wisdom of the crowd, where the amalgamation of multiple models often results in an improved prediction. In the implementation approach, we selected the SVM, XGB, and RF as the first-level models. Each model lent its distinct predictive capabilities to the ensemble with the aim of augmenting the overall performance of the resultant prediction. We conducted a thorough examination of various meta-models for fluid overload prediction to assess their efficacy in integrating the first-level models’ predictions. Choque fuzzy integral fusion, voting classifier, Naïve Bayse for different distributions, LR, RF, and a three-layer multi-layer perceptron network were analyzed. The Gaussian Naïve Bayse model was finally selected as the meta-model.

To find the optimal hyperparameters for the machine learning models of the discriminator, we conducted a broad search encompassing the most influential parameters across various models. Supplemental Table 1 represents the hyperparameters and their values that were analyzed using a grid search strategy to identify the optimal hyperparameters. The area under the receiver operating characteristic (AUROC) was the target performance metric while applying 5-fold crossvalidation. Given the limited number of positive fluid overload cases, choosing accuracy as the target metric for optimization can potentially be misleading. However, AUROC encapsulates a more holistic view of the model’s classification performance and is not biased by the imbalanced class distribution. Hence, a model with a higher AUROC can lead to a more proficient model in classifying fluid overload by maintaining the balance between sensitivity and specificity metrics. Furthermore, we implemented the backward elimination method to identify the most influential variables to predict the presence of fluid overload and subsequently build a logistic regression based on them. Six variables (age, sex, sepsis, SOFA, APACHE II, bicarbonate) were determined based on their significance and the Wald test. Eventually, the performance of the developed models was assessed using AUROC, accuracy, sensitivity, specificity, positive predictive value (PPV), and negative predictive value (NPV).

## Results

### Data characteristics

The distribution and similarities between two synthetically generated datasets and the original dataset were investigated with several analyses. The Jensen-Shannon divergence (JSD) was applied to gauge the similarity between each synthetic dataset and the original one. JSD, a method characterized by its finiteness, symmetry, and smoothing, leverages the principle of information entropy to quantify the divergence between two distributions. For the dataset created using SMOTE, the JSD was 0.10, while the JSD for the dataset produced by the CTGAN model was higher at 0.68. This substantial discrepancy primarily arises from the differing distributions between the synthetic dataset associated with the CTGAN model and the original dataset (most notably that the CTGAN model’s synthetic dataset had a 43.9% prevalence of positive cases compared to a 10.8% occurrence in the original dataset). The Bhattacharyya Distance, a measure useful in comparing datasets with non-normal distributions, further quantified the similarity between the two datasets. The Bhattacharyya Distance was calculated based on the Bhattacharyya coefficient to measure the amount of overlap between the two distributions. Generally, a smaller Bhattacharyya Distance denotes a larger overlap, implying a higher similarity between the distributions, whereas a larger distance indicates less overlap and more divergence. Mann-Whitney U test was also applied. We then used the Benjamini-Hochberg (BH) procedure to manage the false discovery rate [32]. This statistical approach entailed arranging the p-values in ascending order, assigning them respective ranks, and subsequently contrasting each with a computed threshold. Supplemental Table 2 presents the distribution analysis derived from pairwise comparisons of the employed variables. It should be noted that the label distributions of the datasets were entirely different, and we did not set any clinically acceptable range for the synthetically generated data and allowed the CTGAN model to determine this independently.

Figure 2 presents a box plot illustrating the distribution of APACHE II at 24 hours, the SOFA score at 24 hours, MRCICU at 24 hours, and the fluid balance at 24 hours for both the original and CTGAN’s synthetically generated data. These four variables were previously determined to be the most influential on the machine learning models developed [25]. The distributions of the SOFA score, MRC-ICU, and fluid balance in both datasets were relatively similar whereas the APACHE II score values tended to be higher for the synthetic dataset. Given the differences in the label distributions between these two datasets, APACHE II was deemed by the CTGAN model as a significant variable in positive fluid overload cases.

**Fig. 2.**
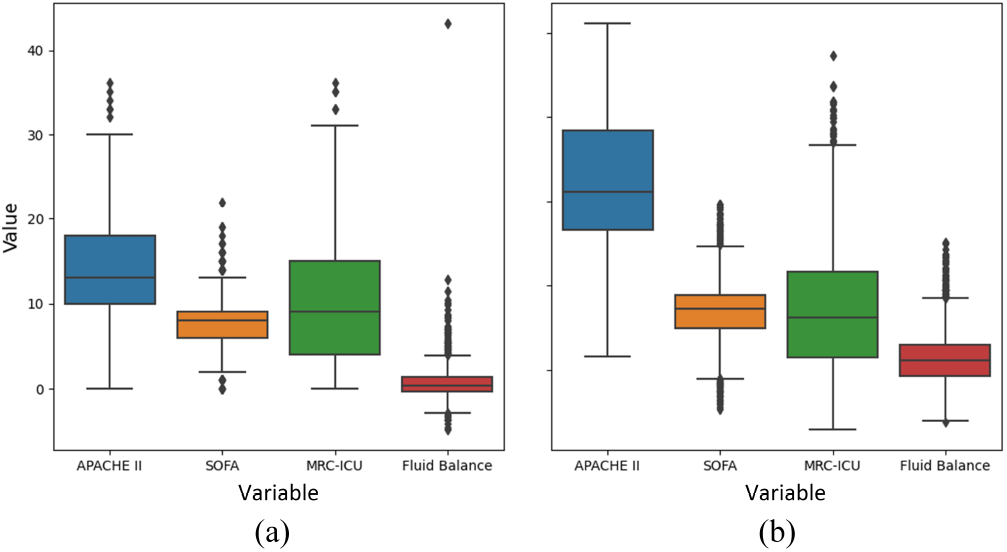
Box plot of four influential variables to predict fluid overload for two different datasets. a) Original dataset. b) CTGAN’s dataset.

Figure 3 illustrates the t-distributed stochastic neighbor embedding (t-SNE) algorithm visualization [33] of the original and CTGAN synthetically generated datasets with the perplexity of 30. The t-SNE aims to find a projection of the data into a lower-dimensional space that preserves the structure of the data to the greatest extent possible. Specifically, the primary goal of t-SNE is to conserve the local structure of the data, implying that points in close proximity within the high-dimensional space are also situated closely within the corresponding low-dimensional space. The t-SNE plot for the original dataset, which exhibited a significant imbalance in terms of label distribution, revealed that the data does not segregate into distinct clusters. This pattern persisted in CTGAN’s synthetically generated dataset, where data points were dispersed along a diagonal line. This observation indicated that predicting fluid overload in ICU patients is an intricate task.

**Fig. 3.**
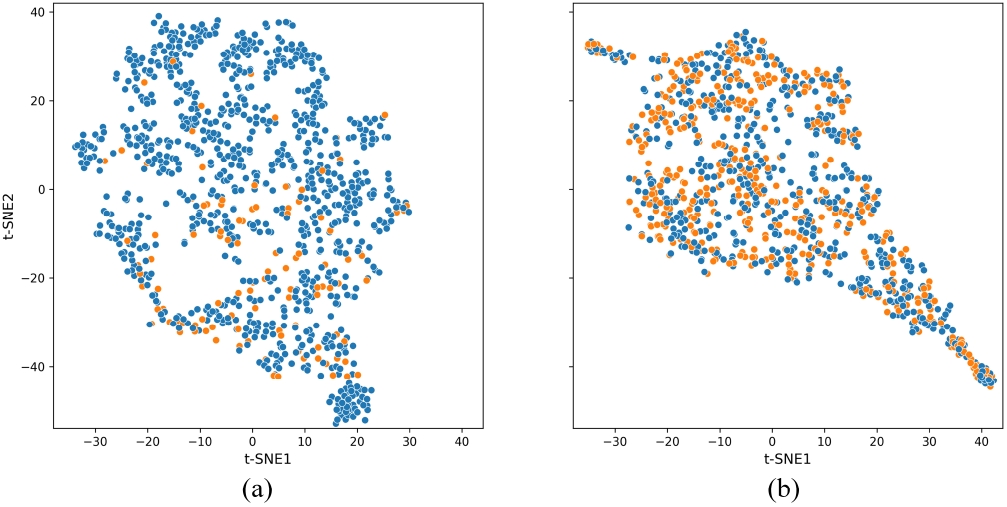
The t-SNE plot of positive/negative fluid overload cases for different datasets. a) Original dataset. b) CTGAN’s dataset.

### Predictive models

Three distinct scenarios were considered in training and evaluating various machine learning models: original data, oversampling the original data using SMOTE, and integrating oversampled original data with CTGAN synthetically generated data. Because of the randomized nature of the imputation, machine learning models, and synthetic data generation, we repeated the training and evaluation of all the scenarios ten times and employed 5-fold crossvalidation. Table 1 summarizes the models’ performance under the three scenarios. Findings from the prior study carried out by Sikora et al. [25] have also been incorporated into the table for comparison.

**Table 1.**
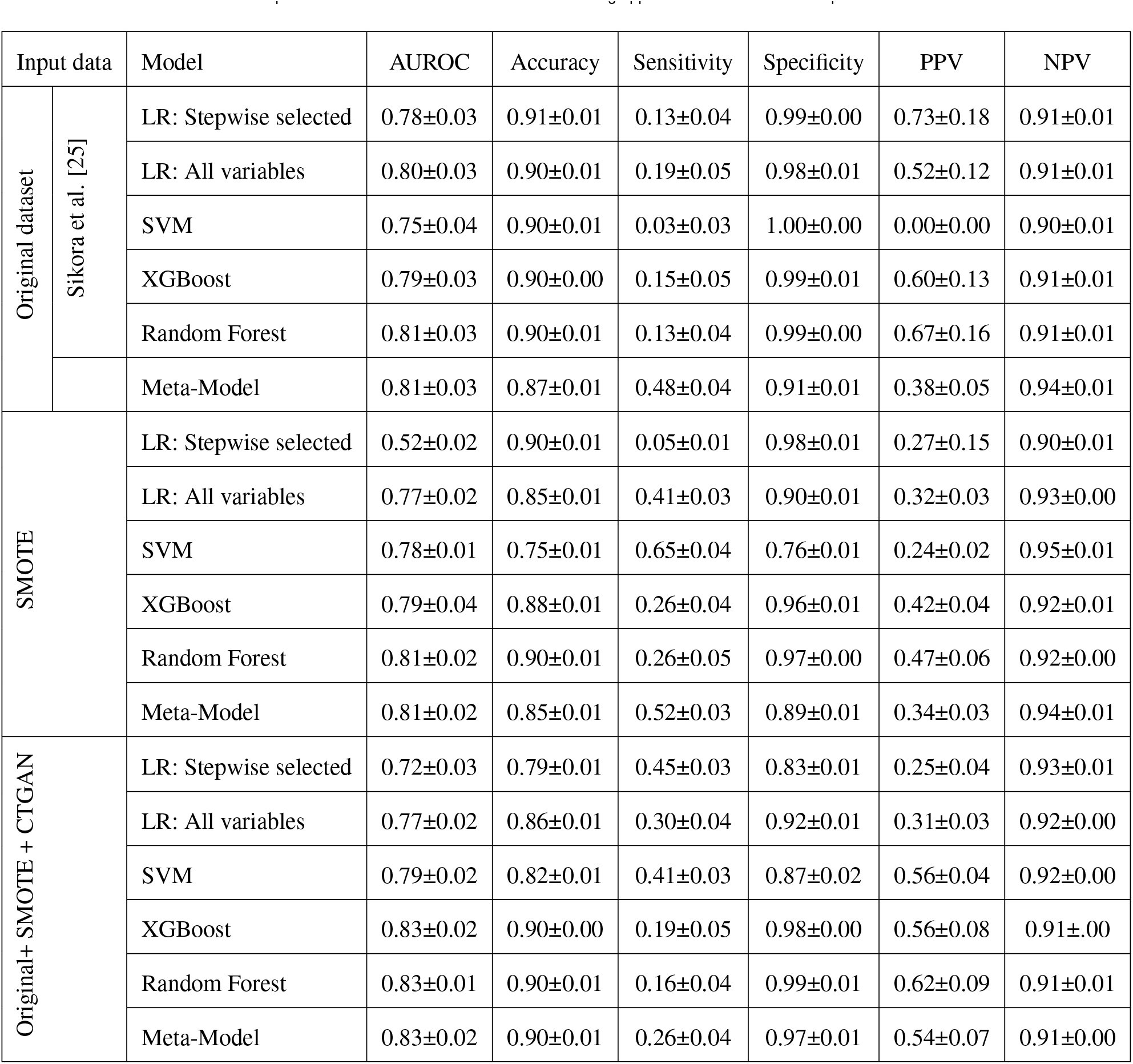
The performance metrics of various machine learning approaches and model development scenarios.

| Input data |  | Model | AUROC | Accuracy | Sensitivity | Specificity | PPV | NPV |
| --- | --- | --- | --- | --- | --- | --- | --- | --- |
| Original dataset | Sikora et al. [25] | LR: Stepwise selected | 0.78±0.03 | 0.91±0.01 | 0.13±0.04 | 0.99±0.00 | 0.73±0.18 | 0.91±0.01 |
|  |  | LR: All variables | 0.80±0.03 | 0.90±0.01 | 0.19±0.05 | 0.98±0.01 | 0.52±0.12 | 0.91±0.01 |
|  |  | SVM | 0.75±0.04 | 0.90±0.01 | 0.03±0.03 | 1.00±0.00 | 0.00±0.00 | 0.90±0.01 |
|  |  | XGBoost | 0.79±0.03 | 0.90±0.00 | 0.15±0.05 | 0.99±0.01 | 0.60±0.13 | 0.91±0.01 |
|  |  | Random Forest | 0.81±0.03 | 0.90±0.01 | 0.13±0.04 | 0.99±0.00 | 0.67±0.16 | 0.91±0.01 |
|  |  | Meta-Model | 0.81±0.03 | 0.87±0.01 | 0.48±0.04 | 0.91±0.01 | 0.38±0.05 | 0.94±0.01 |
| SMOTE |  | LR: Stepwise selected | 0.52±0.02 | 0.90±0.01 | 0.05±0.01 | 0.98±0.01 | 0.27±0.15 | 0.90±0.01 |
|  |  | LR: All variables | 0.77±0.02 | 0.85±0.01 | 0.41±0.03 | 0.90±0.01 | 0.32±0.03 | 0.93±0.00 |
|  |  | SVM | 0.78±0.01 | 0.75±0.01 | 0.65±0.04 | 0.76±0.01 | 0.24±0.02 | 0.95±0.01 |
|  |  | XGBoost | 0.79±0.04 | 0.88±0.01 | 0.26±0.04 | 0.96±0.01 | 0.42±0.04 | 0.92±0.01 |
|  |  | Random Forest | 0.81±0.02 | 0.90±0.01 | 0.26±0.05 | 0.97±0.00 | 0.47±0.06 | 0.92±0.00 |
|  |  | Meta-Model | 0.81±0.02 | 0.85±0.01 | 0.52±0.03 | 0.89±0.01 | 0.34±0.03 | 0.94±0.01 |
| Original+ SMOTE + CTGAN |  | LR: Stepwise selected | 0.72±0.03 | 0.79±0.01 | 0.45±0.03 | 0.83±0.01 | 0.25±0.04 | 0.93±0.01 |
|  |  | LR: All variables | 0.77±0.02 | 0.86±0.01 | 0.30±0.04 | 0.92±0.01 | 0.31±0.03 | 0.92±0.00 |
|  |  | SVM | 0.79±0.02 | 0.82±0.01 | 0.41±0.03 | 0.87±0.02 | 0.56±0.04 | 0.92±0.00 |
|  |  | XGBoost | 0.83±0.02 | 0.90±0.00 | 0.19±0.05 | 0.98±0.00 | 0.56±0.08 | 0.91±0.00 |
|  |  | Random Forest | 0.83±0.01 | 0.90±0.01 | 0.16±0.04 | 0.99±0.01 | 0.62±0.09 | 0.91±0.01 |
|  |  | Meta-Model | 0.83±0.02 | 0.90±0.01 | 0.26±0.04 | 0.97±0.01 | 0.54±0.07 | 0.91±0.00 |

Focusing on the original data scenario, the LR model when constructed on all variables outperformed both the SVM and XGB models in terms of AUROC and sensitivity, scoring 0.80 and 0.19, respectively. While the RF model achieved the highest AUROC of 0.81 and a sensitivity of 0.13, the proposed meta-model succeeded in more than tripling the sensitivity (0.48) while still maintaining the highest AUROC. Turning to the SMOTE scenario, the SVM, XGB, and RF models yielded AUROCs of 0.78, 0.79, and 0.81, and sensitivities of 0.65, 0.26, and 0.26, respectively. The metamodel adeptly managed a trade-off among various performance metrics: it maintained the RF’s AUROC while exhibiting enhanced capability in identifying positive cases, achieving a sensitivity of 0.52. In the CTGAN scenario, even though the LR model built on all variables and the SVM achieved a higher sensitivity than XGB and RF (0.30 and 0.41 compared to 0.19 and 0.16 respectively), the latter models represented a higher AUROC of 0.83. The developed metamodel represented a superior performance by attaining 0.83 AUROC and 0.26 sensitivity.

Generally, the LR model exhibited a lower AUROC using stepwise selected variables compared to the LR model built on all variables across all the scenarios. The addition of a synthetically generated dataset to the training scheme improved the sensitivity of the LR models. Oversampling the dataset using SMOTE enhanced both the AUROC and sensitivity of the SVM, XGB, RF, and meta-model. This increment continued for the AUROC, reaching a peak of 0.83 for the meta-model when CTGAN’s synthetic data was added and representing a more balanced trade-off among the other performance metrics.

Figure 4 illustrates the AUROC curve including a 95% confidence interval for the five developed models, which were trained and validated using all the original and synthetically generated datasets. The meta-model, RF, and XGB demonstrated a similar curve pattern and higher AUROC. While the SVM model yielded a higher AUROC than the LR model built on all variables, the latter exhibited a higher true positive rate at lower false positive values. The LR model built on stepwise selected variables showed a significant difference, although maintaining a curve pattern fairly comparable to the others.

**Fig. 4.**
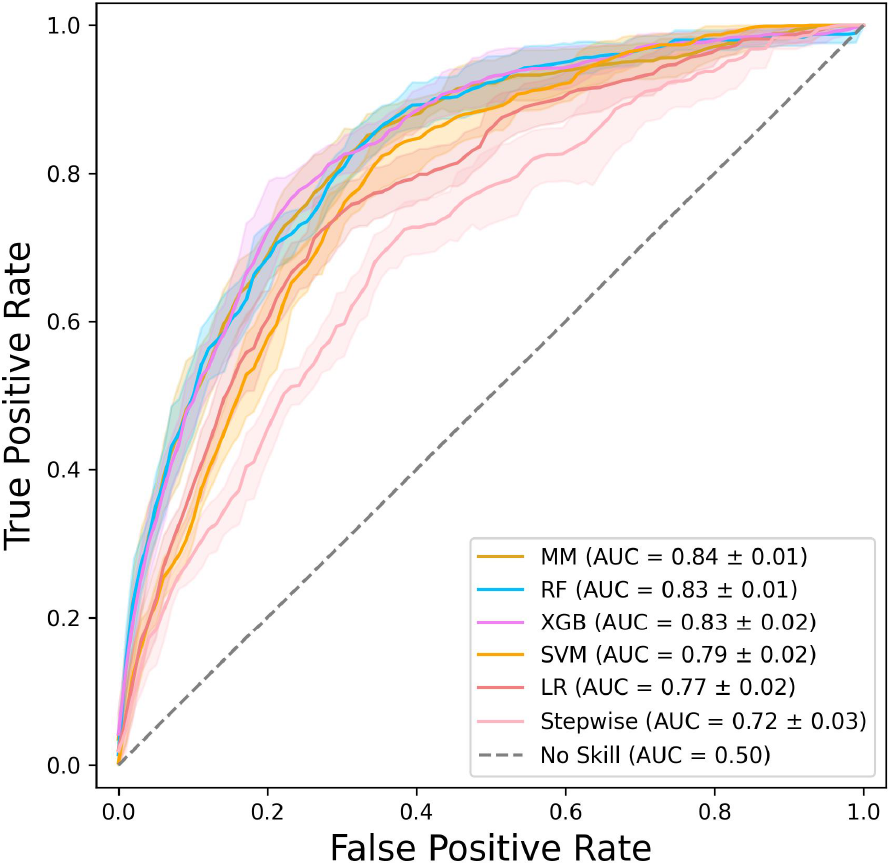
ROC curves of different machine learning models trained on the Original+SMOTE+CTGAN dataset.

Figure 5 shows the hierarchical SHapley Additive exPlanations (SHAP) plot of the meta-model, elucidating the contribution of each variable toward the final prediction [34]. The output of the XGB model emerged as the most influential variable for the developed meta-model using all datasets. Diagnosis of sepsis and rhabdomyolysis, along with the maximum amount of chloride, were ranked as the top three influential variables for the XGB and RF models in predicting fluid overload. For the SVM model, on the other hand, the APACHE II score, MRC-ICU score, and length of stay were deemed the most impactful variables. Supplemental Figures 1 and 2 illustrate the average absolute SHAP value for the inputs of different models for the original dataset and all datasets training scenarios.

**Fig. 5.**
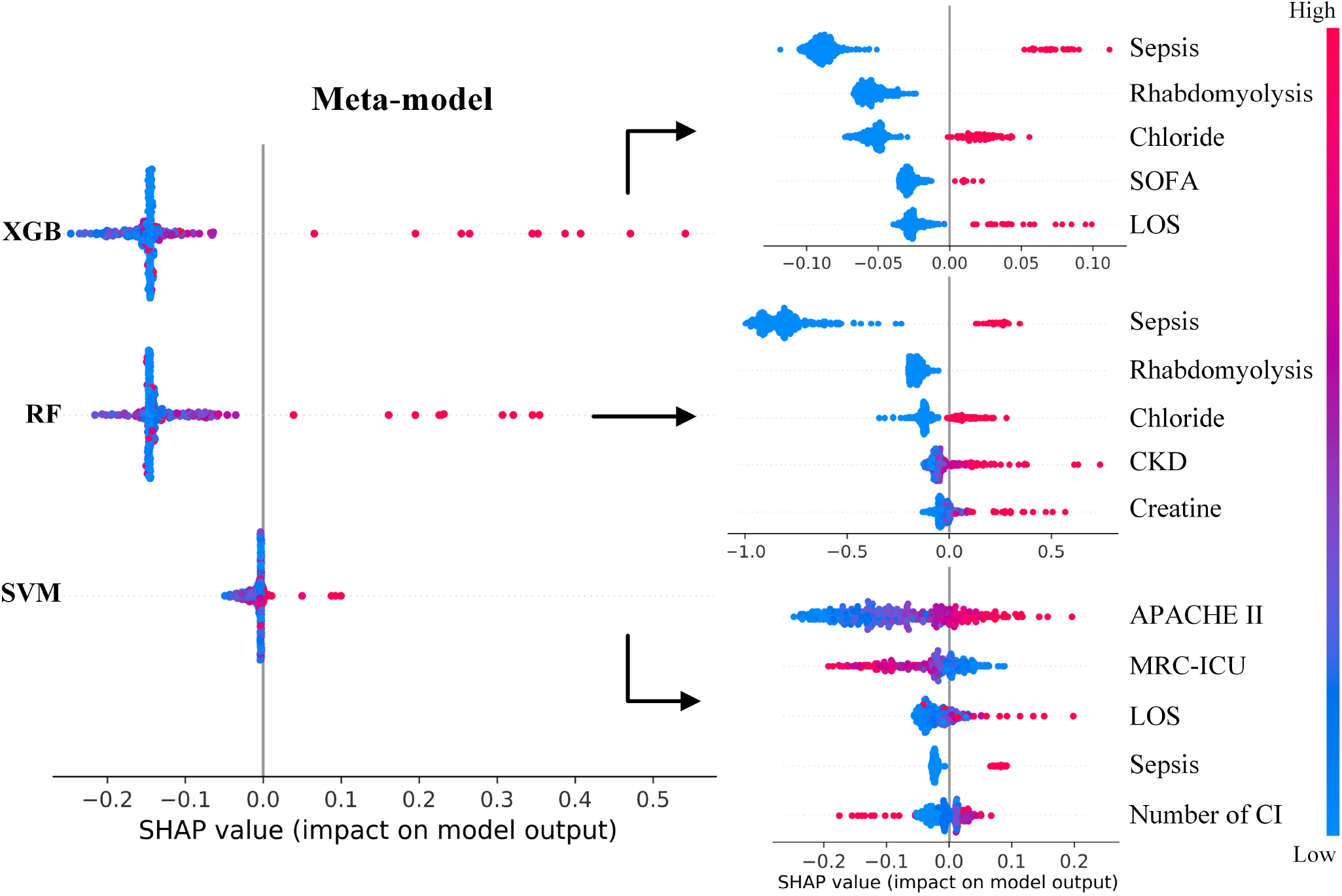
SHAP plot of the meta-model and its first-tier machine learning algorithms trained on the Original+SMOTE+CTGAN dataset.

## Discussion

In the first study to employ conditional GANs to improve irregular temporal modeling for ICU medication data, we found that generating more training data in combination with employing a meta-learner can increase the performance of machine learning algorithms in predicting fluid overload in ICU patients. This study demonstrates the potential of generating synthetic data for the advancement of machine learning methods to predict imbalanced outcomes in critically ill patients. Predicting fluid overload is a demanding task given that the associated data lends itself more towards not having fluid overload than having it and incorporates patientspecific and comprehensive medication data. As the application of machine learning to address the challenge of managing ICU medication has received limited attention, this evaluation may have ultimate applications beyond fluid overload prediction.

The developed meta-learner trained on the combination of synthetically generated and original datasets was able to predict fluid overload at 72 hours with an AUROC of 0.83. While identifying the presence of fluid overload is easy to calculate (simply subtracting the ‘outs’ from the ‘ins’ on a fluid balance flowsheet), predicting its presence such that it could be prevented or its degree lessened through proactive intervention is a clinically complex challenge that has been the subject of far less study [4 20-23 28 35-37]. Given that many critically ill patients require aggressive fluid resuscitation (e.g., in the case of shock) or intravenous medications with a larger volume burden (e.g., intravenous antibiotics), it is unsurprising that fluid overload is a common ICU occurrence. However, its frequency can belie its relationship to adverse ICU events including acute kidney injury, mechanical ventilation, and prolonged length of stay and moreover is potentially mistakenly viewed as ‘unavoidable’ in the context of critical illness [20 38-40]. This assumption is worthy of further investigation given that there are likely several case scenarios within the four phases of volume optimization (Resuscitation, Optimization, Stabilization, and dEresuscitation (ROSE Model). Indeed, while the express goal of the resuscitation phase is to increase circulating volume, the shift to optimization and stabilization requires a nuanced approach that may support either reduced overall volume intake (to limit future fluid overload) or even volume removal. As such, there are likely several case scenarios: (1) a patient requires volume resuscitation in the first 24 hours (or so) of the ICU stay and the clinical scenario (e.g., end organ dysfunction, hemodynamic status, IV medication requirements) makes fluid overload unavoidable, (2) a patient requires volume resuscitation but the clinical scenario allows for euvolemia to be more quickly achieved, (3) a patient did not specifically require volume resuscitation but did require IV medications that resulting in unavoidable fluid overload, or (4) a patient did not specifically require volume resuscitation but did require IV medications that resulting in avoidable fluid overload. The power of predictive algorithms specifically is within scenarios 2 and 4, where with alerts, a clinician could potentially make changes to the patient’s regimen (e.g., adding a diuretic, concentrating IV fluids) that can avoid fluid overload and its potential sequelae. Prior to the advent of machine learning technology, this ability to parse nuanced clinical scenarios was largely left to the expertise and intuition of the bedside clinician; however, the possibility exists that a machine learning-based algorithm could provide meaningful predictions of fluid overload risk in addition to complication (e.g., acute kidney injury) risk in conjunction with potential interventions to help guide the clinician.

Despite the promising potential of machine learning algorithms, one challenge that persists is the limited availability of high-quality data for training models. Acquiring sufficient ICU patient and medication data is crucial for developing practical algorithms; however, collecting, organizing, and structuring these data for fluid overload risk prediction can be time-consuming and costly. Particularly, organizing the granularity of medication data needed (timing, volume, concentration, drug identity, clinical condition, etc.) is a considerable task. Furthermore, the highly imbalanced nature of the available data makes it increasingly challenging for an AI model to discern patterns in cases of positive fluid overload. Synthetic data can provide a valuable solution for addressing the class imbalance in training machine learning models [15 19 41]. Generating synthetic instances of underrepresented classes allows the model to learn more robustly, improving its performance on minority classes and ultimately yielding better generalization. This diversity enhances the model’s understanding of the feature space, aiding it to capture complex patterns more accurately. Synthetic data can also help mitigate privacy concerns and ethical issues in healthcare, as it does not involve using actual patient information [16]. Therefore, we investigated the impact of integrating synthetically generated data in the training phase to assess the effectiveness of different machine learning models in predicting fluid overload in ICU patients. We considered different machine learning approaches and developed a meta-learner as a second-tier learning model that aggregates the predictions of multiple base learners. Overall, by adding more synthetic data to the training phase, the performance of SVM, XGB, and RF was increased. The increase in the sensitivity of these models was higher when the SMOTE data were added while other metrics generally improved by adding the CTGAN dataset. The meta-model was able to make a better trade-off between the performance metrics by increasing both the sensitivity and AUROC across all training scenarios. The dataset generated using CTGAN demonstrated a reasonably equitable label distribution. While the APACHE II score in this dataset exhibited the most significant deviation in distribution compared to the original dataset, the SVM model, which was superior in detecting positive fluid overload cases, considered the APACHE II score as the most influential input variable. The CTGAN model, however, generated other important predictors, notably the MRC-ICU, with a distribution nearly identical to that of the original dataset. Thus, the patterns uncovered using this model placed emphasis on the relationship among the variables instead of generating a singularly accurate variable.

The interoperability and generalizability of developed machine learning models to enhance their understandability, accountability, and robustness were emphasized throughout the study. The complexity introduced by a multi-tiered model can make the interpretation of the machine learning algorithm more challenging. Therefore, we provided a hierarchical SHAP plot for the meta-model to better comprehend how the meta-learner assesses different models and the significance of various variables for first-tier models (Figure 5). The analyses of the interoperability of the meta-model revealed that diagnosing sepsis for a patient had the highest influence on the prediction of fluid overload (Supplemental Figures 1 and 2). Additionally, we augmented the generalizability of the models by incorporating more synthetic data into the training phase, replicating the entire workflow from data imputation to model training ten times, performing cross-validation, conducting an extensive hyperparameter search, and developing a meta-learner.

This study has several limitations: as previously discussed, the original dataset was relatively small in sample size and bias may exist regarding which patients had fluid data available. While the predictors of fluid overload were chosen based on previous evidence, it likely does not represent a complete list of elements that could predict fluid overload. Second, variables were chosen at the 24 hour time point, which while useful for making decisions on the second or third day of the ICU stay, likely fails to capture the highly dynamic nature of ICU patient clinical courses. Finally, the performance of the developed machine learning models is expected to be influenced by the configuration and size of the synthetically generated dataset. Modifying the hyperparameters or adjusting the quantity of synthetic data used for training the machine learning models may lead to different results. Despite these limitations, this methodology marks the first time CGANs have been applied to unstructured ICU medication data and may be transferable to other prediction problems in the ICU domain, particularly those that involve medication therapy.

## Conclusion

This study showcased the potential for synthetic data generation in combination with meta-learner development applied to ICU medication therapy to augment datasets generated from ICU patients to improve prediction performance using the scenario of fluid overload prediction. These methodologies may have utility in a variety of other ICU prediction tasks.

## Data Availability

All data produced in the present study are available upon reasonable request to the authors

## Acknowledgments

Data acquisition was supported by NC TraCS, funded by Grant Number UL1TR002489 from the National Center for Advancing Translations Sciences at the National Institutes of Health, and Data Analytics at the University of North Carolina Medical Center Department of Pharmacy.

