## Supplemental for "Improving irregular temporal modeling by integrating synthetic data to the electronic medical record using conditional GANs: a case study of fluid overload prediction in the intensive care unit"

Table 1. The search space of different hyperparameters of the machine learning models.

| Model | Parameter | Range |
|---|---|---|
| SVM | C | {0.2, 0.5, 0.8, 1.5, 3, 5, 10, 25, 50} |
| | Kernel | {linear, poly, rbf, sigmoid} |
| | Degree | {2, 3, 4, 8} |
| XGBoost | Number of trees | {100, 250, 500} |
| | Maximum depth | {5, 7, 12, 15} |
| | Learning rate | {0.01, 0.1} |
| | Subsample columns by tree | {0.6, 0.8, 1} |
| | Gamma | {0, 0.1, 1} |
| | Scale positive ratio | {1, 1.1, 1.2, 1.5, 2} |
| Random Forest | Number of trees | {100, 150, 200, 300, 500, 1000, 1500, 3000} |
| | Maximum depth | {5, 8, 10, 12, 14, 18} |
| | Minimum samples leaf | {1, 2, 4} |
| | Minimum samples split | {2, 4, 5, 10} |
| | Class weight | {1:1, 1:2, 1:5} |

Table 2. Distribution analysis of the variables used in the development of the machine learning models. The values from two synthetically generated datasets are compared with those from the original dataset.

| **Variables** | **Bhattacharyya Distance** | | **Mann-Whitney U test** | |
|---|---|---|---|---|
| | **SMOTE** | **CTGAN** | **SMOTE** | **CTGAN** |
| Sex | 0.0964 | 0.1345 | <0.0001 | <0.0001 |
| Age | 0.0613 | 0.0785 | <0.0001 | 0.2247 |
| ICU type | 0.0377 | 0.0785 | <0.0001 | 0.0004 |
| APACHE II at 24 hours | 0.1134 | 0.1757 | <0.0001 | <0.0001 |
| SOFA at 24 hours | 0.0760 | 0.0785 | <0.0001 | <0.0001 |
| MRC-ICU at 24 hours | 0.0130 | 0.0785 | <0.0001 | <0.0001 |
| AKI at 24 hours | 0.1134 | 0.0317 | 0.0003 | 0.1191 |
| Use of CRRT at 24 hours | 0.0371 | 0.0476 | 0.0051 | <0.0001 |
| Any CRRT | 0.0371 | 0.0785 | <0.0001 | <0.0001 |
| Mechanical ventilation before 24 hours | 0.0371 | 0.0596 | <0.0001 | <0.0001 |
| Any mechanical ventilation | 0.0384 | 0.0476 | <0.0001 | <0.0001 |
| Creatinine | 0.0371 | 0.0615 | <0.0001 | <0.0001 |
| Sodium | 0.0613 | 0.0785 | <0.0001 | <0.0001 |
| Chloride | 0.0064 | 0.0317 | <0.0001 | <0.0001 |
| Potassium | 0.0377 | 0.0615 | <0.0001 | 0.0002 |
| Bicarbonate | 0.1513 | 0.0785 | <0.0001 | <0.0001 |
| Vasopressor use in first 24 hours | 0.0371 | 0.0785 | 0.0004 | <0.0001 |
| Use of continuous infusion | 0.0049 | 0.0785 | 0.8084 | <0.0001 |
| Number of continuous infusions | 0.0613 | 0.0785 | 0.0117 | <0.0001 |
| Fluid balance at 24 hours | 0.0732 | 0.0785 | <0.0001 | <0.0001 |
| Length of ICU stay | 0.1134 | 0.2393 | <0.0001 | <0.0001 |
| Other (coded diagnosis) | 0.0444 | 0.0785 | <0.0001 | 0.5825 |
| Trauma | 0.0371 | 0.0476 | 0.0001 | 0.1191 |
| Sepsis | 0.0371 | 0.0785 | 0.0065 | <0.0001 |
| Hepatic | 0.0371 | 0.0785 | 0.0023 | <0.0001 |
| Cardiac | 0.0613 | 0.0476 | <0.0001 | 0.2374 |
| Pulmonary | 0.0613 | 0.0655 | <0.0001 | <0.0001 |
| Pancreatitis | 0.0371 | 0.0785 | 0.0384 | <0.0001 |
| pulmonary arterial hypertension | 0.0233 | 0.0316 | 0.4529 | <0.0001 |
| Rhabdomyolysis | <0.0001 | <0.0001 | 1.0000 | <0.0001 |
| Cirrhosis | <0.0001 | <0.0001 | 1.0000 | 0.0101 |
| Heart failure | 0.0377 | 0.0476 | <0.0001 | 0.0450 |
| Chronic kidney disease | 0.0384 | 0.0785 | <0.0001 | <0.0001 |

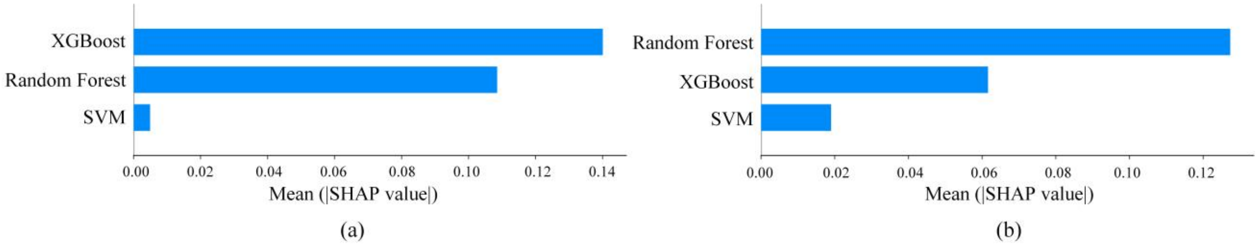


Figure 1. The average absolute SHAP values of each first-tier model of the developed meta-learner. a) Trained on the Original+SMOTE+CTGAN datasets. b) Trained on original datasets.

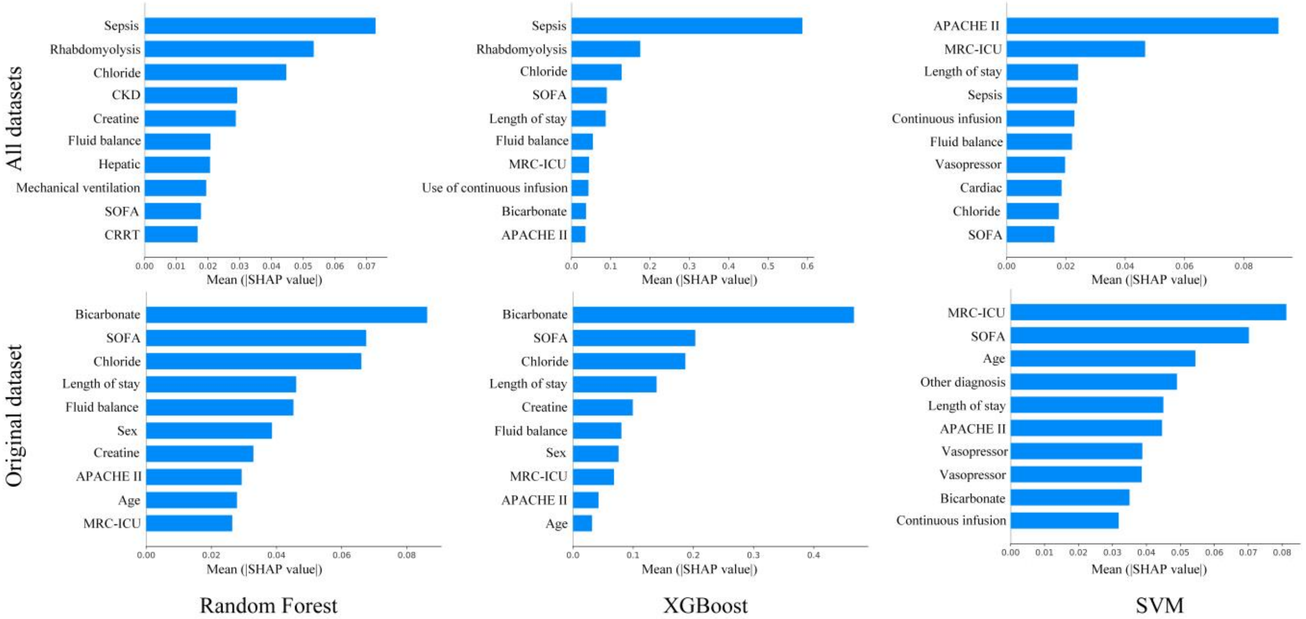


Figure 2. The average absolute SHAP values for each variable across different machine learning models trained on the Original+SMOTE+CTGAN datasets and trained on only the original datasets.
